# Radiomics-based functional outcome prediction in acute single subcortical infarction

**DOI:** 10.1101/2024.07.08.24310116

**Authors:** Tang Yang, Yue Ming, Yajun Cheng, Shuai Jiang, Yuying Yan, Yi Hu, Chen Ye, Ruosu Pan, Jiayu Sun, Bo Wu

## Abstract

**Background:** Clinical outcomes vary considerably among patients with acute single subcortical infarction (SSI). We aimed to construct a model incorporating radiomic features and clinical factors to predict functional outcomes in patients with acute SSI.

**Methods:** We enrolled patients who experienced acute SSI within 14 days of stroke onset and randomly divided them into training (n=118) and test (n=30) cohorts. Unfavorable functional outcome was defined as a modified Rankin Scale score >1 at 3 months. We extracted and selected radiomics features from baseline diffusion-weighted imaging and perfusion-weighted imaging to develop a radiomics model. Multivariate logistic regression was performed to construct a clinical model using clinical factors and imaging features. Finally, a combined model was built using both clinical and radiomics features. Receiver operating characteristic curves were used to evaluate the discriminatory ability of these models.

**Results:** The radiomics model, encompassing 13 radiomics features, exhibited good predictive performance for unfavorable functional outcomes with area under the curve (AUC) values of 0.774 and 0.824 in the training and test cohorts, respectively. The combined model, which included clinical factors (early neurological deterioration, hypertension, baseline National Institutes of Health Stroke Scale score, infarct volume, and summary cerebral small vessel disease score) and radiomics features, improved performance in the training (AUC = 0.915) and test (AUC = 0.846) cohorts.

**Conclusions:** The clinical-radiomics model provided improved accuracy for the prognostic prediction of SSI, which may help clinicians in the decision-making process and improve long-term outcomes in patients with SSI.

## Introduction

Single subcortical infarction (SSI) within the territory of a perforating artery accounts for approximately a quarter of all ischemic strokes.^1^ Most single subcortical infarctions are due to an intrinsic small vessel pathology with a small proportion due to embolism or parent large artery or branch artery atheroma.^2^ SSI usually has a small infarction volume and is considered to have a favorable outcome in comparison with other stroke subtypes.^3^ However, the prognosis of SSI considerably depends on risk factors and demographic characteristics,^3^ as well as the different etiological mechanisms of SSI, and the presence of early neurological deterioration (END).^4,5^ As a result, the prediction of clinical outcomes at an early stage would be beneficial for patients with SSI, as clinicians could select individualized therapeutic strategies for these patients.

With rapid advances in machine learning, radiomics has emerged as a computer-aided process. By transforming neuroimaging data into numerous quantitative features (e.g., shape, intensity or texture), radiomic data are expected to help neurologists diagnose stroke, predict early outcome, and evaluate long-term prognoses.^6^ Perfusion-weighted imaging (PWI) allows for very sensitive detection of perfusion deficits in SSI patients within the first hours after the onset of symptoms despite their small volume.^7,8^ Some studies have attempted to apply clinical and radiomic data to predict functional outcomes after acute ischemic stroke, and their results suggested that diffusion-weighted imaging (DWI) radiomics could be applied as a prognostic indicator,^9,10^ but little is known about whether the radiomic features attracted from both DWI and PWI could indicate the poor prognosis in SSI patients.

Therefore, we aimed to investigate whether the radiomic model could predict the functional outcomes at 3 months in patients with SSI and to develop a prediction model incorporating both clinical factors and radiomics features extracted from DWI and PWI images.

## Methods

### Clinical Data

We reviewed data from the prospectively collected acute SSI database of the Department of Neurology at West China Hospital between November 2018 and January 2023. The inclusion criteria were as follows: (1) a first-ever stroke with compatible SSI in the unilateral LSA territory (basal ganglia, internal capsule, and corona radiata) confirmed by DWI; and (2) DWI and PWI examination within 14 days of onset. The exclusion criteria were as follows: (1) patients who had ≥ 50% stenosis of the ipsilateral middle cerebral artery or internal carotid artery on computed tomography angiography (CTA) or magnetic resonance angiography (MRA); (2) patients with non-atherosclerotic vasculopathies, such as moyamoya disease, dissection, or vasculitis; (3) patients with high-risk factors for cardioembolism (e.g., atrial fibrillation, patent foramen ovale, valvular heart disease, or infective endocarditis) identified by transthoracic echocardiography and 24-h electrocardiographic or Holter monitoring; and (4) patients with poor image quality. This study was approved by the Biomedical Research Ethics Committee of West China Hospital (No. 2020 [324]). Signed informed consent was obtained from all participants or their legally authorized representatives. Patients were randomly dichotomized into the training (n = 118) and test (n = 30) cohorts at a ratio of 8:2.

Baseline data, including age, sex, smoking status, drinking status, hypertension, diabetes mellitus, hyperlipidemia, infarct volume, and onset to magnetic resonance imaging (MRI) time were recorded. The National Institutes of Health Stroke Scale (NIHSS) was used to assess the severity of neurological deficits. END was defined as an increase of ≥2 points in the total NIHSS score or ≥1 point in the motor NIHSS score in comparison with the best neurological status (including symptom fluctuations) in the period of 7 days after stroke onset.^11^ Functional outcomes were evaluated by the modified Rankin Scale (mRS) score at 3 months and classified as an excellent outcome (mRS score ≤1) or an unfavorable outcome (mRS score >1).

### MRI Acquisition

MRI was performed on a 3.0 Tesla Siemens Trio MRI scanner (Siemens Medical Systems, Erlangen, Germany) with a 32-channel head coil. The MRI protocol included T1-weighted, T2-weighted, fluid-attenuated inversion recovery imaging, DWI, susceptibility-weighted imaging, 3D time-of-flight MRA, and dynamic susceptibility contrast-PWI. PWI scans were obtained using a gradient-echo echo-planar imaging sequence, and perfusion images were acquired (repetition time=500 ms, echo time=32 ms, flip=90, field-of-view=220 × 220 mm, 19 slices with a slice thickness of 5 mm, matrix size=128 × 128, scan time=83 s) after intravenous injection (injection speed, 4 ml/s) of 0.2 mmol/kg of a gadolinium-based contrast agent (Magnevist; Schering, Berlin, Germany). The imaging parameters of the other sequences were reported in our previous studies.^12^

### Assessment of Cerebral Small Vessel Disease MRI Markers

The four MRI markers of cerebral small vessel disease (CSVD) were defined according to the standards for reporting vascular changes on neuroimaging 2 (STRIVE-2) criteria.^2^ The total burden of CSVD was expressed by a CSVD compound score, which ranges from 0 to 4. The methods used to assess CSVD imaging markers and count summary CSVD scores have been described in full previously.^13^

### Imaging Analysis

PWI images were processed using Siemens Syngo.via VB40 (Siemens Healthineers, Erlangen, Germany) and the MR Neurology package with the arterial input function. The perfusion maps of cerebral blood flow (CBF), cerebral blood volume (CBV), mean transit time (MTT) and time to peak (TTP) and the DWI images were all resampled to a uniform resolution (1 mm × 1 mm × 1 mm). Then, the CBF, CBV, MTT, and TTP maps were co-registered with the DWI images, respectively.

### Lesion Segmentation and Radiomics Feature Extraction

The process of the radiomics analysis included lesion segmentation, feature extraction, feature selection, and model building (Figure 1). Infarction lesions were manually delineated along a high signal on DWI slice by slice with ITK SNAP (http://www.itksnap.org) by two experienced investigators (T.Y. and Y.J.C.) blinded to the clinical data. The interclass correlation coefficient was 0.89 for lesion segmentation on DWI. The region of interest (ROI) (the hyperintense area of SSI) on DWI was registered to the corresponding perfusion maps (CBF/CBV/MTT/TTP) in SSI region.

**Figure 1.**
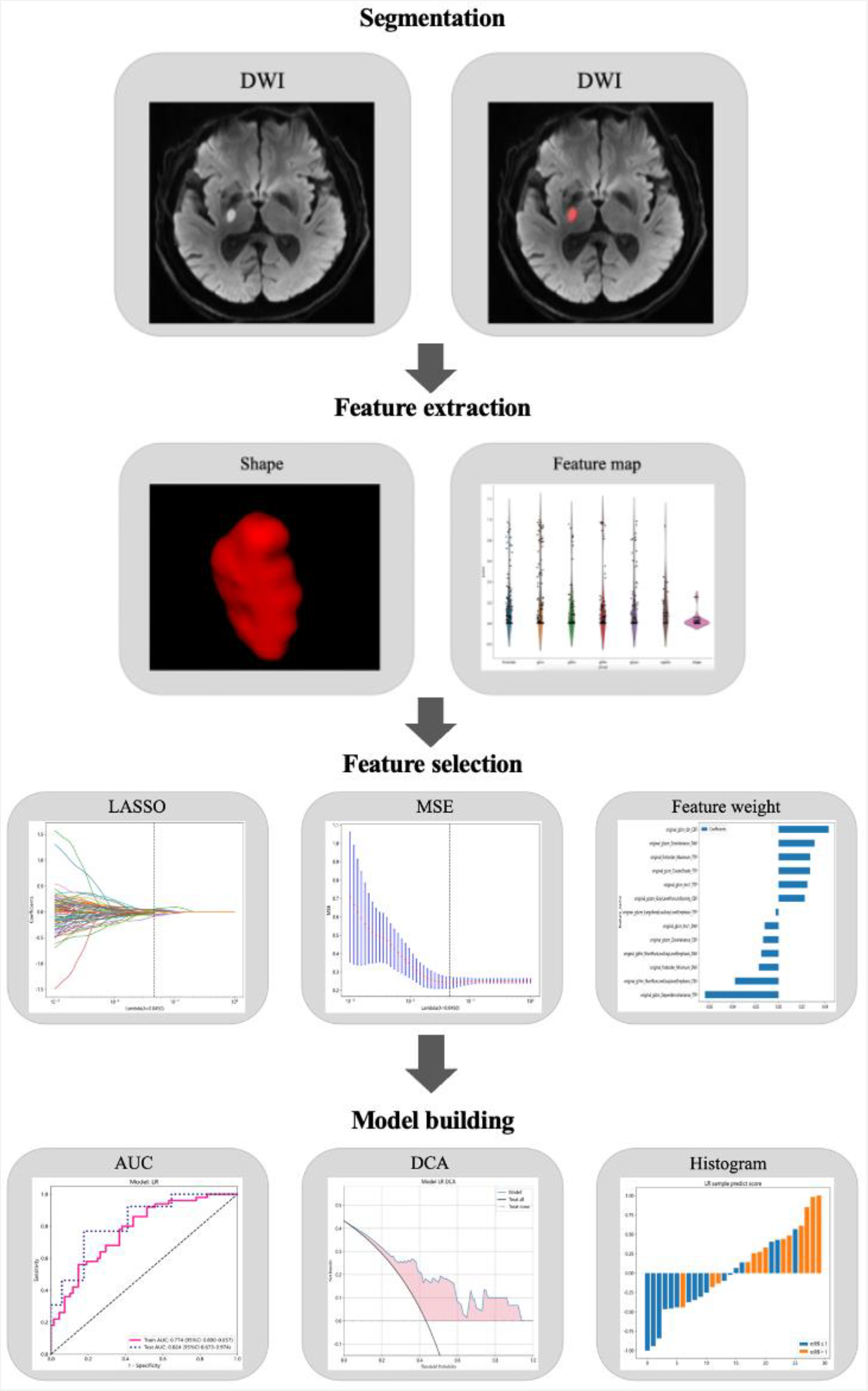
The procedure of radiomic analysis.

Radiomics features were extracted based on the Pyradiomics Python package. In total, 535 features were extracted for each ROI. The radiomics features included seven categories: 90 first-order features; 70 shape features; 70 gray level dependence matrix (GLDM) features; 120 gray level co-occurrence matrix (GLCM) features; 80 gray level run length matrix (GLRLM) features; 80 gray level size zone matrix (GLSZM) features; and 25 neighboring gray tone difference matrix (NGTDM) features.

### Feature Selection and Radiomics Model Building

In the step of feature selection, we conducted a t test or Mann-Whitney U test and feature screening for all radiomic features. Only radiomic features with P < 0.05 were retained. Then, we used Spearman’s correlation coefficient to calculate the correlation between features to eliminate redundant features. Next, the least absolute shrinkage and selection operator (LASSO) regression model was applied to select outcome-relevant features, and a ten-fold cross validation was used to select optimal features. The radiomics score was obtained by a linear combination of retained features weighted by the LASSO algorithm. After LASSO feature screening, we input the final features into the logistic regression model for radiomics model building.

### Clinical and Combined Prediction Models

The features (early neurological deterioration, hypertension, baseline NIHSS score, infarct volume, and summary small vessel disease score) used for building the clinical model were selected by univariate logistic regression analysis (P values<0.05). The multivariate logistic regression analysis was implemented to construct a clinical prediction model. Finally, the combined model was established by combining clinical features and radiomic features.

### Statistical analysis

Quantitative variables were presented as mean ± standard deviation (SD) or as median with interquartile range. The Shapiro-Wilk test was used to check for a normal distribution. Quantitative variables were assessed using Student’s t test or the Mann-Whitney U test, while categorical variables were analyzed using the chi-squared test or Fisher’s exact test. Receiver operating characteristic (ROC) curves were drawn to evaluate the discrimination ability of the above models. The calibration curve and Hosmer–Lemeshow test were applied to evaluate the calibration efficiency of the three models. Decision curve analysis (DCA) was used to determine the clinical utility of these models on the basis of calculating the net benefits at each threshold probability. Statistical analyses were conducted using the R software package (version 4.3.1). Statistical significance was determined by a two-sided p value < 0.05.

## Results

### Patient characteristics

A total of 148 patients were included. The mean age was 55.93±10.19 years, and 117 (79.1%) patients were male. The baseline demographic, clinical, and radiological data are presented in Table 1. We analyzed the differences in baseline data between the excellent (mRS ≤ 1) and unfavorable (mRS > 1) functional outcome groups. Patients with unfavorable outcomes were more likely to have hypertension, END, a higher NIHSS score, a larger infarct volume, and a higher summary CSVD score. In the multivariate logistic regression analysis, these five variables also showed statistically significant differences between the two groups (hypertension: OR 2.766, 95%CI [1.057, 7.754], P 0.043; END: OR 3.319, 95%CI [1.256, 9.136], P 0.017; NIHSS score: OR 1.623, 95%CI [1.364, 1.983], P <0.001; infarct volume: OR 1.403, 95%CI [1.096, 1.833], P 0.009; summary CSVD score: OR 1.491, 95%CI [1.042, 2.195], P 0.034) (Table 1).

**Table 1.** Univariate and multivariate logistic regression analysis of risk factors.

|  | mRS ≤ 1 (n=85) | mRS > 1 (n=63) | P | Multivariate OR<br>(95%CI) | P |
| --- | --- | --- | --- | --- | --- |
| Age (years) | 54.99±10.08 | 57.19±10.28 | 0.195 |  |  |
| Male sex | 67 (78.8%) | 50 (79.4%) | 1.000 |  |  |
| Smoking | 46 (54.1%) | 36 (57.1%) | 0.842 |  |  |
| Drinking | 29 (34.1%) | 28 (44.4%) | 0.269 |  |  |
| Hypertension | 43 (50.6%) | 45 (71.4%) | 0.017 | 2.766<br>(1.057, 7.754) | 0.043 |
| Diabetes mellitus | 26 (30.6%) | 20 (31.7%) | 1.000 |  |  |
| Hyperlipidemia | 21 (24.7%) | 19 (30.2%) | 0.581 |  |  |
| END | 12 (14.1%) | 28 (44.4%) | <0.001 | 3.319<br>(1.256, 9.136) | 0.017 |
| Onset to MRI time<br>(days) | 5 (4-7) | 5 (4-8) | 0.983 |  |  |
| NIHSS score | 2 (2-3) | 6 (3.5-8) | <0.001 | 1.623<br>(1.364, 1.983) | <0.001 |
| Infarct volume | 1.00 (0.53,<br>1.68) | 1.73 (1.01, 2.86) | <0.001 | 1.403<br>(1.096, 1.833) | 0.009 |
| ≥1 Lacunes | 39 (45.9%) | 34 (54.0%) | 0.420 |  |  |
| ≥1 CMBs | 27 (31.8%) | 30 (47.6%) | 0.074 |  |  |
| Moderate to severe EPVSs | 41 (48.2%) | 42 (66.7%) | 0.039 |  |  |
| WMH | 25 (29.4%) | 23 (36.5%) | 0.463 |  |  |
| Summary CSVD score | 1 (0-3) | 2 (1-3) | 0.045 | 1.491<br>(1.042, 2.195) | 0.034 |

mRS, modified Rankin Scale; END, early neurological deterioration; MRI, magnetic resonance imaging; NIHSS, National Institutes of Health Stroke Scale; CMB, cerebral microbleeds; EPVS, enlarged perivascular spaces; WMH, white matter hyperintensity; CSVD, cerebral small vessel disease; OR, odds ratio; CI confidence interval.

### Performance of the Three Prediction Models

After the LASSO algorithm was applied, 13 features, including one feature on CBV, three features on CBF, four features on DWI, and five features on TTP, were finally selected and used to construct the radiomics model. The distribution of the 13 features is presented in Figure 2. The radiomics model had AUC values of 0.774 [95% confidence interval (CI) 0.691-0.857] in the training cohort and 0.824 (95% CI 0.673-0.974) in the test cohort (Table 2). The DCA for the radiomics model is shown in Figure 3A. The decision curve demonstrated that using the radiomic model to predict outcomes in patients with SSI leads to greater benefit. In addition, the calibration curves suggested that the favorable predictive performance was satisfactorily consistent with the ideal curve in the training cohort (Figure 3B).

**Figure 2.**
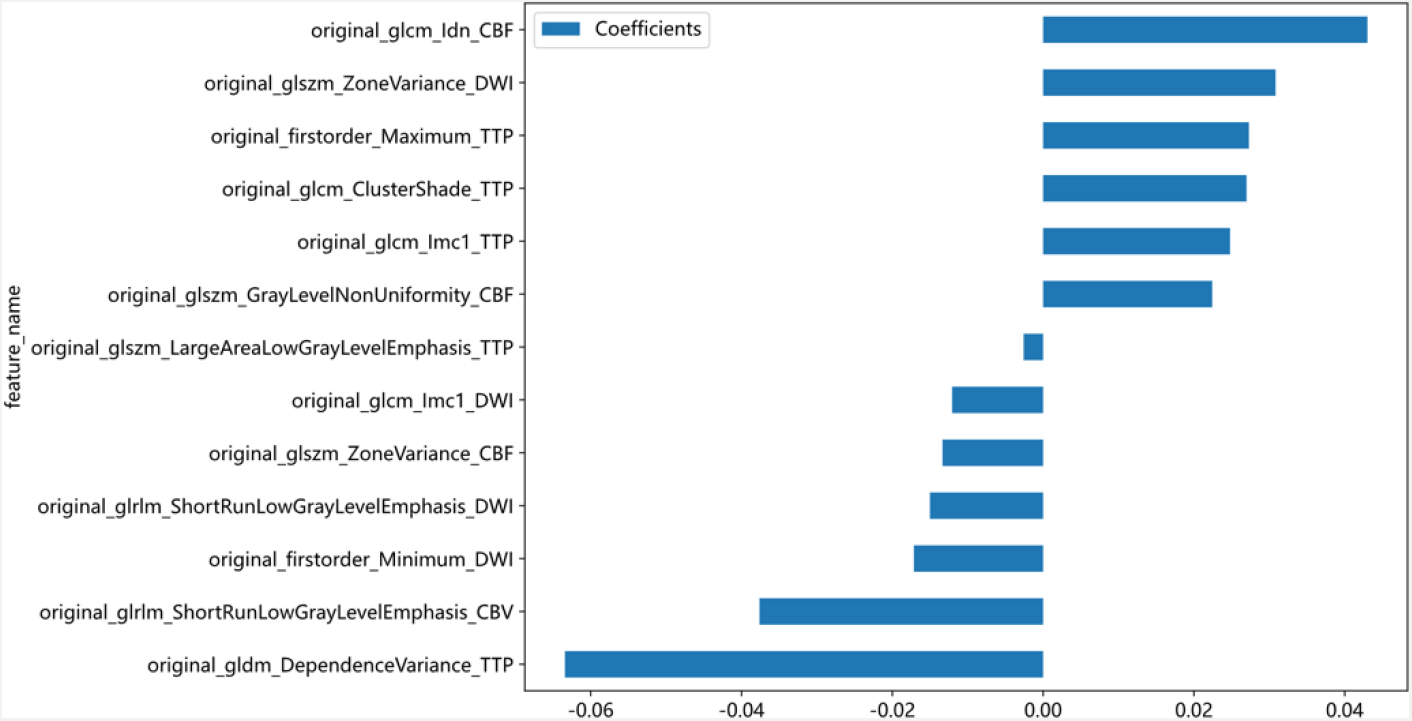
Feature name and distribution.

**Figure 3.**
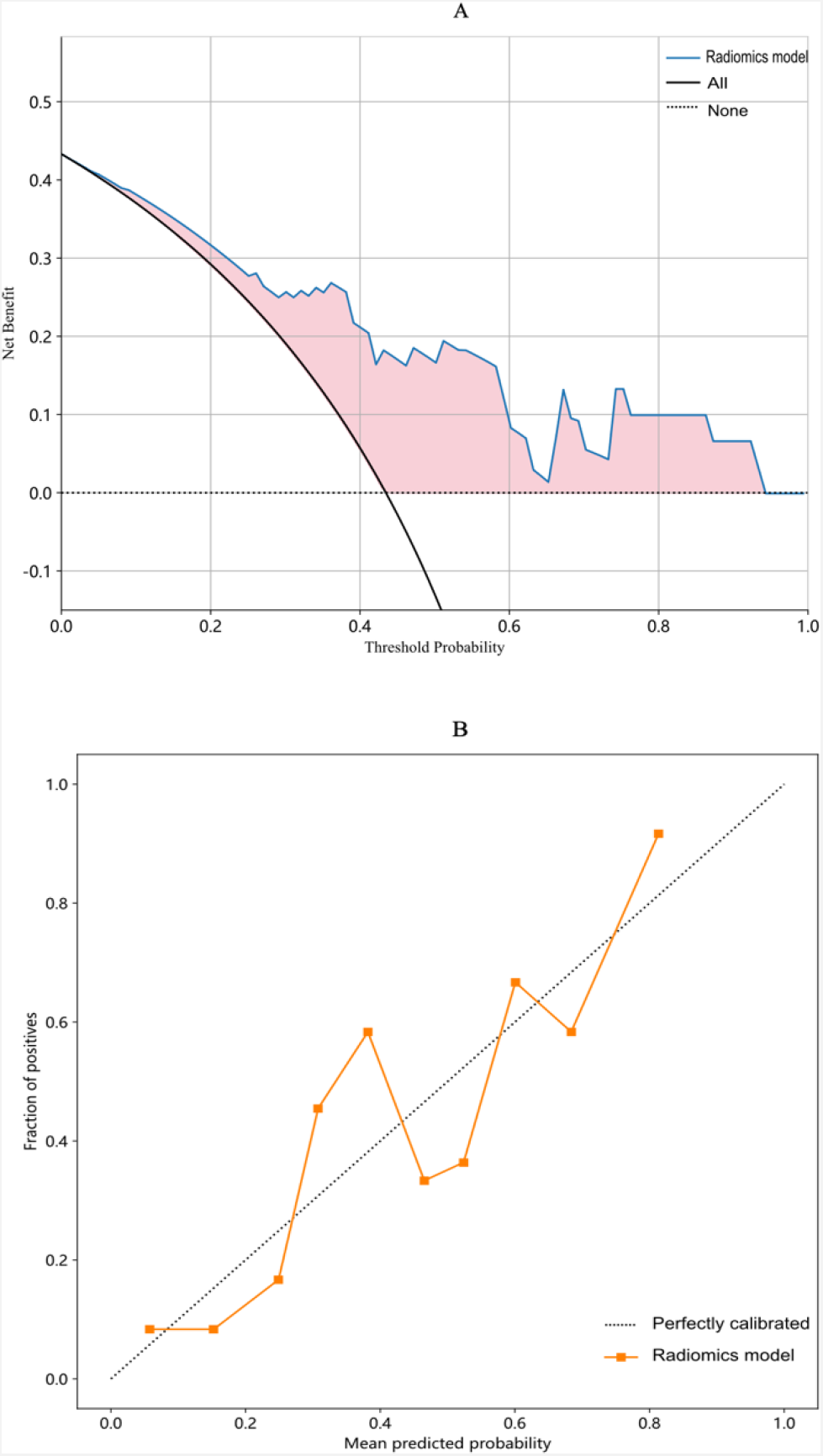
Decision curve and calibration curve analysis of the radiomics model. (A) Decision curve of the radiomic model. (B) Calibration curve for the radiomic model.

**Table 2.** Predictive performance of three models in the training and testing cohorts.

| Model | Training cohort (n=118) |  |  | Test cohort (n=30) |  |  |
| --- | --- | --- | --- | --- | --- | --- |
|  | AUC (95% CI) | Sensitivity | Specificity | AUC (95% CI) | Sensitivity | Specificity |
| Clinical model | 0.897<br>(0.843-0.952) | 0.760 | 0.882 | 0.787<br>(0.617-0.958) | 0.615 | 0.824 |
| Radiomics model | 0.774<br>(0.691-0.857) | 0.840 | 0.559 | 0.824<br>(0.673-0.974) | 0.692 | 0.824 |
| Combined model | 0.915<br>(0.867-0.963) | 0.860 | 0.779 | 0.846<br>(0.693-0.999) | 0.615 | 0.941 |

AUC, area under the curve; CI, confidence interval.

According to the results of the multivariate logistic regression, a clinical model was constructed, which exhibited an AUC value of 0.897 (95% CI 0.843-0.952) in the training cohort and 0.787 (95% CI 0.617-0.958) in the test cohort. The combined model showed greater predictive performance than the radiomics model and clinical model in both cohorts (Figure 4).

**Figure 4.**
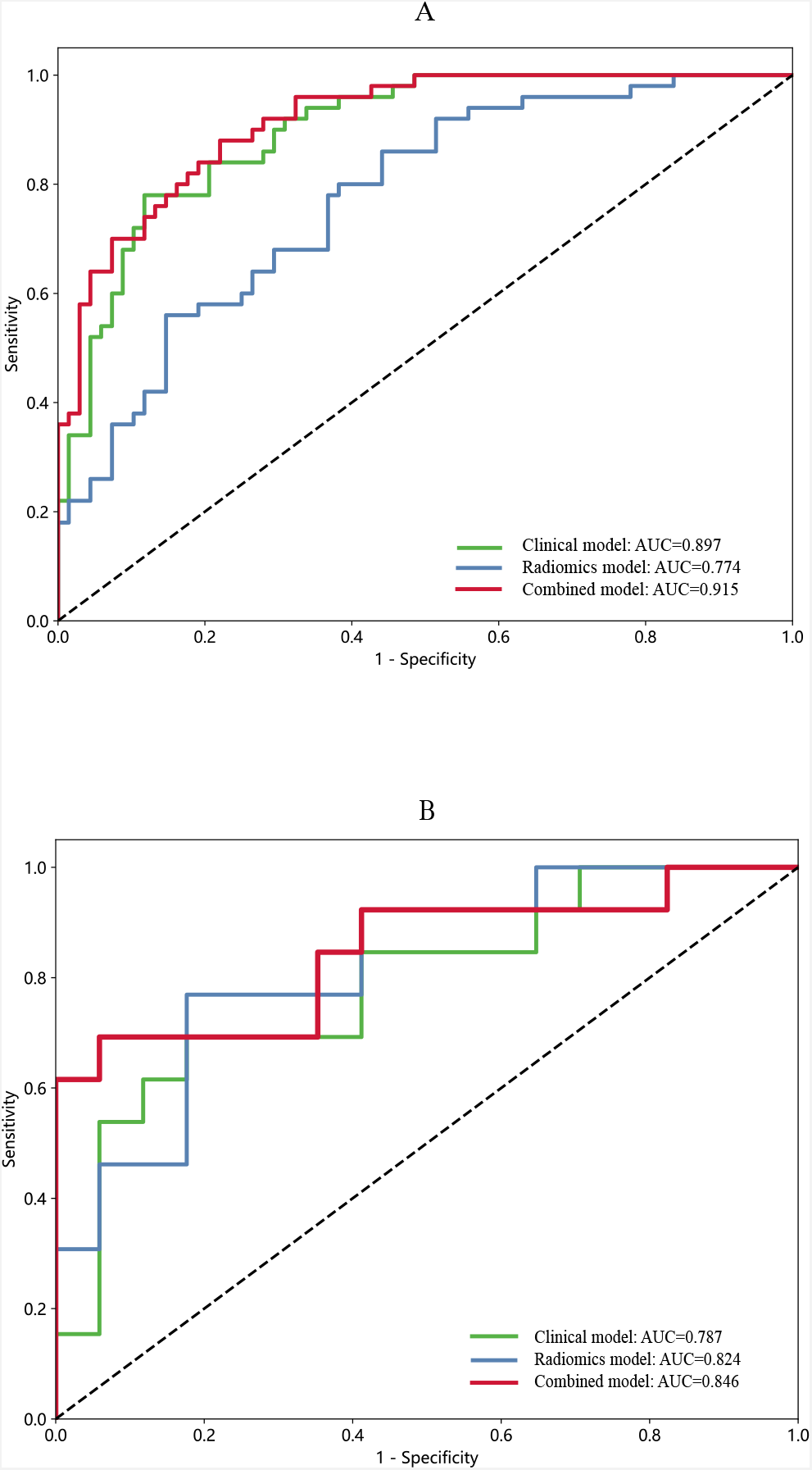
ROC curves of the radiomics model, clinical model, and clinical-radiomics model in the training (A) and testing (B) cohorts. ROC, receiver operating characteristic.

## Discussion

This study demonstrated that the novel prediction model incorporating clinical and radiomic features had improved performance in predicting functional outcomes in patients with SSI. To our knowledge, the present study is the first radiomics-based study to investigate the functional outcomes of acute SSI individuals. As a result, clinicians could obtain more details and predict the prognosis of SSI patients earlier by using the clinical-radiomics model.

The functional outcomes of patients with SSI varied, and those with excellent outcomes at 3 months accounted for 57.4% of our study. Predictors of prognosis might be potential therapeutic targets of SSI. During the past few years, radiomics has shown promise for a variety of applications in stroke, especially in the prediction of long-term outcomes. Wang et al.^10^ combined risk factors (age, NIHSS score at 24 hours post-admission, hemorrhage) and DWI-based radiomics scores to develop a clinical-radiomics model for the prediction of 3-month outcomes in patients with acute ischemic stroke. Their study showed that the AUC values were 0.80 in the training cohort and 0.73 in the validation cohort. A recent study conducted by Zhou et al.^9^ also established a clinical-radiomics model to predict clinical outcomes at 6 months after symptom onset in acute ischemic stroke patients, with AUCs of 0.868 and 0.890 in the training and validation cohorts, respectively. However, they only extracted radiomics features from DWI and apparent diffusion coefficient (ADC) images, which might limit the predictive performance of the model. Another study^14^ revealed that the use of radiomics score (from ADC and CBF maps) and clinical features could predict favorable clinical outcomes in acute ischemic stroke patients at 7 days and 3 months. The results indicated that the radiomics model had played a satisfactory role of complement of the clinical model. Our study included more clinical information such as infarct volume, CSVD markers, and END, and we also extracted more radiomics features from the perfusion maps of CBV, CBF, MTT, and TTP. In the present study, the combined model showed strong predictive value with AUCs of 0.897 and 0.787 in the training and test cohorts, separately.

Radiomics is a promising noninvasive and quantitative tool for prognosis prediction that involves extracting features from computed tomography (CT) or MRI images.^15,16^ Among the 13 selected features in this study, two radiomics features were first-order features and eleven radiomics features were texture features. First-order features indicate the characteristics of imaging exhibited by voxels alone, while texture features indicate other aspects of the image, such as the relationships to the adjacent voxel, linear scales, and voxel blocks.^17^ The original_gldm_DependenceVariance feature had the highest weight coefficient among these 13 features, which represented the heterogeneity or variability of images. Higher values are associated with greater heterogeneity of infarcts. These radiomics features could show changes in the cerebral microstructure, which cannot be quantitatively identified by the naked eye.

However, it is not comprehensive to predict clinical outcomes by using only the radiomics features of infarction lesions. We found that hypertension, END, NIHSS score, infarct volume, and summary CSVD score were independent indicators of clinical outcome at three months in patients with SSI. Our results were consistent with those of previous studies investigating the prognosis of SSI patients.^18–21^ As shown in Figure 2, a novel clinical-radiomics model was constructed combining these independent risk factors and radiomics features, which had improved AUC values to 0.915 in the training cohort and 0.846 in the test cohort. In comparison with prior studies, we applied a radiomics approach to quantify the image data after segmentation of the SSI lesions, achieving greater individualized prediction of functional outcomes at 3 months.

There were some limitations in the current study. First, the sample size was relatively small, and all patients came from one center. Second, the established clinical-radiomics model lacked external validation, and multicenter studies are required to validate the results of our study. Third, since the study was conducted in China, the generalizability of the findings to other populations may be limited. In addition, lesions were manually segmented instead of fully automated, but the reproducibility was excellent.

## Conclusions

In conclusion, our findings suggested that the combined model outperformed individual clinical or radiomics model in predicting the 3-month functional outcomes of SSI patients, which provided new insights into the prognostic prediction of SSI. With advanced imaging and artificial intelligence technology, radiomics methodology might aid clinicians in the decision-making process and improve the long-term outcomes in patients with SSI.

## Data Availability

The data that support the findings of this study are available from the corresponding author upon reasonable request.

## Acknowledgments

None.

## Sources of Funding

This work was supported by grants from the National Key R&D Program of China (2023YFC2506603), the National Natural Science Foundation of China (82071320, 82271328, 82301661, and 82371322), the China Postdoctoral Science Foundation (2022M712249), the Post-Doctor Research Project, West China Hospital, Sichuan University (2023HXBH007), and the 1.3.5 project for disciplines of excellence, Clinical Research Incubation Project, West China Hospital, Sichuan University (2020HXFH012).

## Disclosures

None.

## Notes

### Competing Interest Statement

The authors have declared no competing interest.

### Author Declarations

This study was approved by the Biomedical Research Ethics Committee of West China Hospital (No. 2020 [324]).

## References

1. Petty GW, Brown RD, Whisnant JP, Sicks JD, O’Fallon WM, Wiebers DO. Ischemic Stroke Subtypes: A Population-Based Study of Functional Outcome, Survival, and Recurrence. Stroke. 2000; 31:1062–1068.

2. Duering M, Biessels GJ, Brodtmann A, Chen C, Cordonnier C, De Leeuw F-E, Debette S, Frayne R, Jouvent E, Rost NS, et al. Neuroimaging standards for research into small vessel disease—advances since 2013. The Lancet Neurology. 2023; 22:602–618.

3. Norrving B. Long-term prognosis after lacunar infarction. The Lancet Neurology. 2003; 2:238–245.

4. Zhang C, Wang Y, Zhao X, Wang D, Liu L, Wang C, Pu Y, Zou X, Du W, Jing J, et al. Distal Single Subcortical Infarction Had a Better Clinical Outcome Compared With Proximal Single Subcortical Infarction. Stroke. 2014; 45:2613–2619.

5. Vynckier J, Maamari B, Grunder L, Goeldlin MB, Meinel TR, Kaesmacher J, Hakim A, Arnold M, Gralla J, Seiffge DJ, et al. Early Neurologic Deterioration in Lacunar Stroke: Clinical and Imaging Predictors and Association With Long-term Outcome. Neurology. 2021; 97: e1437–e1446.

6. Chen Q, Xia T, Zhang M, Xia N, Liu J, Yang Y. Radiomics in Stroke Neuroimaging: Techniques, Applications, and Challenges. Aging Dis. 2021; 12:143–154.

7. Doege CA, Kerskens CM, Romero BI, Brunecker P, Junge-Hülsing J, Pannwitz W von, Müller B, Villringer A. Assessment of Diffusion and Perfusion Deficits in Patients with Small Subcortical Ischemia. American Journal of Neuroradiology. 2003; 24:1355–1363.

8. Förster A, Mürle B, Böhme J, Al-Zghloul M, Kerl HU, Wenz H, Groden C. Perfusion-weighted imaging and dynamic 4D angiograms for the estimation of collateral blood flow in lacunar infarction. J Cereb Blood Flow Metab. 2016; 36:1744–1754.

9. Zhou Y, Wu D, Yan S, Xie Y, Zhang S, Lv W, Qin Y, Liu Y, Liu C, Lu J, et al. Feasibility of a Clinical-Radiomics Model to Predict the Outcomes of Acute Ischemic Stroke. Korean J Radiol. 2022; 23:811.

10. Wang H, Sun Y, Ge Y, Wu P-Y, Lin J, Zhao J, Song B. A Clinical-Radiomics Nomogram for Functional Outcome Predictions in Ischemic Stroke. Neurol Ther. 2021; 10:819–832.

11. Li H, Dai Y, Wu H, Luo L, Wei L, Zhou L, Lin Y, Wang Q, Lu Z. Predictors of Early Neurologic Deterioration in Acute Pontine Infarction. Stroke. 2020;51(2):637–640.

12. Jiang S, Cao T, Yan Y, Yang T, Yuan Y, Deng Q, Wu T, Sun J, Wu S, Hao Z-L, et al. Lenticulostriate artery combined with neuroimaging markers of cerebral small vessel disease differentiate the pathogenesis of recent subcortical infarction. J Cereb Blood Flow Metab. 2021; 41:2105–2115.

13. Yang T, Peng P, Jiang S, Yan Y, Hu Y, Wang H, Ye C, Pan R, Sun J, Wu B. Multiple Hypointense Vessels are Associated with Cognitive Impairment in Patients with Single Subcortical Infarction. Transl Stroke Res. 2023 Dec 5. doi: 10.1007/s12975-023-01206-9.

14. Tang T, Jiao Y, Cui Y, Zhao D, Zhang Y, Wang Z, Meng X, Yin X-D, Yang Y-J, Teng G, et al. Penumbra-based radiomics signature as prognostic biomarkers for thrombolysis of acute ischemic stroke patients: a multicenter cohort study. J Neurol. 2020; 267:1454–1463.

15. Mayerhoefer ME, Materka A, Langs G, Häggström I, Szczypiński P, Gibbs P, Cook G. Introduction to Radiomics. J Nucl Med. 2020; 61:488–495.

16. Quan G, Ban R, Ren J-L, Liu Y, Wang W, Dai S, Yuan T. FLAIR and ADC Image-Based Radiomics Features as Predictive Biomarkers of Unfavorable Outcome in Patients With Acute Ischemic Stroke. Front. Neurosci. 2021; 15:730879.

17. Jiang L, Miao Z, Chen H, Geng W, Yong W, Chen Y-C, Zhang H, Duan S, Yin X, Zhang Z. Radiomics Analysis of Diffusion-Weighted Imaging and Long-Term Unfavorable Outcomes Risk for Acute Stroke. Stroke. 2023; 54:488–498.

18. Shin DW, Gorelick PB, Bae H-J, the Clinical Research Collaboration for Stroke in Korea (CRCS-K) Investigators. Time-dependent shift of the relationship between systolic blood pressure and clinical outcome in acute lacunar stroke. International Journal of Stroke. 2022; 17:400–406.

19. Asdaghi N, Pearce LA, Nakajima M, Field TS, Bazan C, Cermeno F, McClure LA, Anderson DC, Hart RG, Benavente OR. Clinical Correlates of Infarct Shape and Volume in Lacunar Strokes: The Secondary Prevention of Small Subcortical Strokes Trial. Stroke. 2014; 45:2952–2958.

20. Liu X, Li T, Diao S, Cai X, Kong Y, Zhang L, Wang Z, Li R, Zhou Y, Fang Q. The global burden of cerebral small vessel disease related to neurological deficit severity and clinical outcomes of acute ischemic stroke after IV rt-PA treatment. Neurol Sci. 2019; 40:1157–1166.

21. Adams HP Jr, Davis PH, Leira EC, Chang KC, Bendixen BH, Clarke WR, Woolson RF, Hansen MD. Baseline NIH Stroke Scale score strongly predicts outcome after stroke: A report of the Trial of Org 10172 in Acute Stroke Treatment (TOAST). Neurology. 1999; 53:126–131.

